# Progression of biological markers in spinocerebellar ataxia type 3: analysis of longitudinal data from the ESMI cohort

**DOI:** 10.1101/2025.01.30.25321426

**Authors:** Moritz Berger, Hector Garcia-Moreno, Monica Ferreira, Jeannette Hubener-Schmid, Tamara Schaprian, Philipp Wegner, Tim Elter, Kennet Teichmann, Magda M Santana, Marcus Grobe-Einsler, Demet Onder, Berkan Koyak, Sarah Bernsen, Luís Pereira de Almeida, Patrick Silva, Joana Afonso Ribeiro, Inês Cunha, Cristina Gonzalez-Robles, Shamsher Khan, Amanda Heslegrave, Henrik Zetterberg, Manuela Lima, Mafalda Raposo, Ana F. Ferreira, João Vasconcelos, Bart P. van de Warrenburg, Judith van Gaalen, Teije H. van Prooije, Jeroen de Vries, Ludger Schols, Olaf Riess, Matthis Synofzik, Dagmar Timmann, Andreas Thieme, Friedrich Erdlenbruch, Jon Infante, Ana Lara Pelayo, Leire Manrique, Kathrin Reetz, Imis Dogan, Gulin Oz, James M. Joers, Khalaf Bushara, Chiadikaobi Onyike, Michal Povazan, Heike Jacobi, Jeremy D Schmahmann, Eva-Maria Ratai, Matthias Schmid, Paola Giunti, Thomas Klockgether, Jennifer Faber

## Abstract

**Background:** Spinocerebellar ataxia type 3 (SCA3) is an autosomal dominantly inherited adult-onset disease. We aimed to describe longitudinal changes in clinical and biological findings and to identify predictors for clinical progression.

**Methods:** We used data from participants enrolled in the ESMI cohort collected between Nov 09, 2016 and July 18, 2023. The data freeze included data from 14 sites in five European countries and the United States. We assessed ataxia with the Scale for the Assessment and Rating of Ataxia (SARA). We measured disease-specific mutant ataxin-3 protein (ATXN3) and neurofilament light chain (NfL) in plasma and performed MRIs. Data were analysed by regression modelling on a timescale defined by onset. The onset of abnormality of a marker was defined as the time at which its value, as determined by modelling, exceeded the mean ±2 SD of healthy controls. To study responsiveness of markers, we determined the sensitivity to change ratios (SCSs).

**Results:** Data from 291 SCA3 mutation carriers before and after clinical onset and 121 healthy controls were included. NfL levels became abnormal more than 20 years (-21.5 years [95% CI n.d. –9.5]) before onset. The earliest MRI abnormality was volume loss of medulla oblongata (-4.7 years [95% CI n.d. – 3.3]). The responsiveness of markers depended on the disease stage. Across all stages, pons volume had the highest responsiveness with an SCS of 1.35 [95% CI 1.11 – 1.78] exceeding that of SARA (0.99 [95% CI 0.88 – 1.11]). Lower age (p=0.0459) and lower medulla oblongata volume (p<0.0001) were predictors of SARA progression.

**Conclusion:** Our study provides quantitative information on the progression of biological markers in SCA3 mutation carriers before and after onset of ataxia, and allowed the identification of predictors for clinical progression. Our data could prove useful for the design of future clinical trials.

## Introduction

Spinocerebellar ataxia type 3 (SCA3) is the most common autosomal dominantly inherited adult-onset ataxia disease worldwide. SCA3 takes a progressive course and leads to increasing disability and premature death. It is caused by unstable expansions of polyglutamine encoding CAG repeats within the *ATXN3* gene, resulting in the formation of abnormally elongated, misfolded ataxin-3 protein (ATXN3).^1^

Targeted therapies for SCA3 are being developed, and first safety trials of antisense oligonucleotides (ASOs) have been initiated (https://clinicaltrials.gov, NCT05160558, NCT05822908). In the future, preventive intervention in mutation carriers before clinical onset will be a realistic option.^2^ With the advent of disease-modifying treatments for SCA3, there is the need to identify biological markers that are sensitive to disease-related change before and after clinical manifestation. Mutant ATXN3 can be measured at low concentrations in the CSF and plasma of mutation carriers, but is absent in healthy controls.^3,4^ Blood neurofilament light chain (NfL) is an easily accessible, non-specific marker of neurodegeneration.^5^ In cross-sectional studies, NfL was increased in patients and in mutation carriers before onset.^6–10^ In a two-year follow-up study of 19 SCA3 patients, the increased NfL concentrations did not change.^9^ In longitudinal MRI studies of SCA3 mutation carriers, progressive atrophy of the cerebellum, pons, mesencephalon, and cervical spinal cord was observed.^11–14^ In addition, diffusion parameters of cerebellar peduncles, superior longitudinal fasciculus, corona radiata, and medial lemniscus showed increasing abnormalities.^13–15^

The European Spinocerebellar ataxia type 3/Machado-Joseph disease Initiative (ESMI) initiated a longitudinal registry study of SCA3 mutation carriers before and after clinical onset representing a wide spectrum of disease severity. Analysis of cross-sectional clinical, as well as fluid biomarker and MRI volumetric data allowed to draft a data-driven model of disease stages for SCA3.^16^ In the present study, we describe longitudinal changes of mutant ATXN3, NfL and several MRI measures that were abnormal in SCA3 mutation carriers before onset. We focused the analysis on determining stage-specific sensitivity of biological markers and identifying predictors of clinical progression.

## Methods

### Study design and participants

The study population of the ESMI registry study consists of (1) SCA3 mutation carriers before and after onset, (2) persons at risk to carry the SCA3 mutation (first degree relatives of SCA3 patients) who have not been diagnostically tested, and who do not wish to be tested, and (3) healthy controls (including spouses, unrelated persons, and persons at risk who were negatively tested). Following anonymous genetic testing (see: Procedures), persons at risk were assigned either to the SCA3 mutation carrier or healthy control group.

The ESMI registry study is conducted at 14 sites in five European countries and the United States. Participants undergo annual standardized assessments including clinical examination and biosample collection. MRI is performed at 11 sites.

The study was approved by the ethics committees of all contributing centres. At enrolment, informed and written consent following the Declaration of Helsinki was obtained from all study participants. The study protocol is available online (https://ataxia-esmi.eu/study-protocols).

### Procedures

We used the Scale for the Assessment and Rating of Ataxia (SARA)^17^ to assess the presence and severity of ataxia. Manifest ataxia was defined by a score of ≥ 3.^17,18^

Analysis of the CAG repeat length of the *ATXN3* gene was performed at the Department of Medical Genetics of the University of Tübingen (Tübingen, Germany). Determinations were done for 243 mutation carriers and 20 persons at risk who had not been diagnostically tested. Results of the genetic tests in persons at risk were not disclosed to study participants. For 42 SCA3 mutation carriers, from whom no DNA was available, information about CAG repeat lengths was taken from medical records; in eight participants, no information on repeat length was available.

Plasma concentrations of mutant ATXN3 were measured using an ultrasensitive immunoassay based on the SMC® technology.^3^ Plasma concentrations of NfL were determined with the Neurology 4-Plex A assay (N4PA) (Quanterix, Billerica, MA, United States) run on the Simoa HD-X Analyzer™.^7^ Samples were analysed using two different assay lots. For each sample, measurements were performed in split duplicates, and the average values were calculated.

T1- and diffusion weighted MRIs were acquired on Siemens 3T scanners (Siemens Medical Systems, Erlangen, Germany). As imaging biological markers, we calculated 61 brain volumes including brainstem and cerebellar sub-segments and the mean diffusion metrics (fractional anisotropy (FA), medial diffusivity (MD), axial (AD) and radial diffusivity (RD)) of 14 white matter tracts. Details of the MR sequences and imaging analysis as well as a comprehensive list of all studied volumes and white matter tracts are given in the appendix pp 4-5.

### Statistical analysis

Statistical analysis was carried out using R version 4.3.1 (R Core Team 2023: R: A Language and Environment for Statistical Computing, R Foundation for Statistical Computing, Vienna, Austria).

The selection of MRI parameters was based on the group comparison between pre-ataxic SCA3 mutation carriers with a SARA <3 and healthy controls using linear regression models. Details on the statistical methods and test results are given in the appendix pp 6-7.

Age of onset was defined as the reported first occurrence of gait disturbances.^19^ 36 SCA3 mutation carriers had not yet experienced gait disturbances (right-censored individuals). In nine SCA3 mutation carriers with gait disturbance, information on the reported age of onset was missing (left-censored individuals). In these 45 SCA3 mutation carriers, the age of onset was estimated, as described below. Five NfL values, one ATXN3 value, and one pons volume value were excluded as outliers after visual inspection of the data (appendix p 2).

To relate fluid and MRI biomarker data to the time from onset, we applied a conditional multiple imputation approach.^16^ First, censored values of age of onset were imputed fitting a previously published parametric survival model.^20^ For this, the last follow-up visit of each participant was considered and the time from onset for towards each visit date was then calculated respectively. To account for censoring, age of onset was imputed with the conditional expectation for right-censored individuals (accounting for actual age) and with the unconditional expectation for left-censored individuals. Second, SARA score and biological markers were regressed on the (imputed) time from onset using additive mixed regression models with participant-specific random intercepts and a cubic P-spline with six B-spline basis functions and a second-order difference penalty. This two-step procedure was repeatedly applied to 1000 bootstrap samples from the original sample. Final estimates of the spline coefficients and associated variance estimates were then calculated by applying Rubin’s rule.

For regression, NfL concentrations and MRI measures were z-transformed with respect to age and sex. Z-scores of MRI volumes and FA values were inverted, so that higher z-scores indicate increasing abnormality in all measures. Since SARA scores and mutant ATXN3 in healthy controls were close to 0, no z-transformation was performed, and the raw values were used. A Box-Cox transformation with parameter λ= 0.25 was applied to the SARA score to approximate normality.

A biological marker was considered abnormal, if its value, as determined by modelling, exceeded the normal range defined by mean ± 2 SD in healthy controls. Onset of abnormality was defined as not determinable (n.d.) if the intersection between the normal range and the fitted spline function of the upper and lower limit of the 95% CI, respectively, was not reached within the time interval of observations (33 years before to 41 years after onset). Following recently proposed definitions of SCA3 disease stages,^16^ SCA3 mutation carriers were assigned to the carrier stage (SARA < 3 and NfL z-score < 2), biomarker stage (SARA < 3 and NfL z-score ≥ 2), or ataxia stage (SARA ≥ 3).

Responsiveness of SARA and biological markers was assessed by calculating sensitivity to change ratios (SCSs).^21^ To this end, linear mixed regression models with the (imputed) time from onset as the time variable and participant-specific random intercepts were fitted for the entire disease course and each stage (carrier, biomarker, ataxia), respectively. SCSs were then calculated by dividing the estimated slopes of progression by the estimated standard deviation of the slopes. 95% CIs of the SCSs were determined by nonparametric bootstrap based on 1000 samples. Higher SCS values indicate greater sensitivity to change of the respective measure.

For prediction of SARA increase, we applied univariable and multivariable mixed regression models with the Box-Cox transformed SARA as outcome and age, sex, CAG repeat length of the expanded allele and the baseline values of the biological markers as covariates. We tested the effect of these factors on SARA progression by interactions with the time variable (time from onset). The multivariable model was selected by stepwise selection with the Bayesian information criterion including all covariates with p<0.05 in univariable models. We conducted the univariable analysis for the entire disease course and each stage (carrier, biomarker, ataxia), respectively, while we restricted the multivariable model to the entire disease course to ensure a sufficiently large sample size.

## Results

Between Nov 09, 2016, and Jul 18, 2023, we enrolled 419 participants with at least one available biological marker. Seven participants and biomarker data from 39 visits were excluded. A flow chart detailing the reasons is given in the appendix p 2. Eventually, 291 SCA3 mutation carriers and 121 healthy controls were included in the analysis. Among the SCA3 mutation carriers, 55 had no ataxia (SARA < 3), and 236 had ataxia (SARA ≥ 3) at baseline. At baseline, mutant ATXN3 concentrations were available in 97, NfL concentrations in 303, and MRI results in 171 participants. Baseline characteristics of the study participants and the subgroups with available biological markers are given in table 1.

**Table 1:** Baseline characteristics of study participants.

|  | Healthy controls | SCA3 mutation carriers<br>with SARA < 3 | SCA3 mutation carriers<br>with SARA ≥ 3 |
| --- | --- | --- | --- |
| **Total group (n = 412)** |  |  |  |
| Number of participants | 121 (29%) | 55 (13%) | 236 (57%) |
| Women | 71 (59%) | 30 (55%) | 113 (47.9%) |
| Age, years | 44.6 (34.3 – 56.3) | 34.6 (29.1 – 39.9) | 52.2 (45.2 – 60.3) |
| SARA score | 0.0 (0.0 – 0.5) | 1.0 (0.5 – 2.0) | 12.0 (8.5 – 19.0) |
| Length of expanded CAG allele | n.a. | 68 (65 – 71) | 69 (66 – 71) |
| Time from onset, years | n.a. | -12.5 (-16.8 – -0.0) | 10.1 (5.6 – 14.9) |
| **ATXN3 subgroup (n = 97)** |  |  |  |
| Number of participants | 10 (10%) | 14 (14%) | 73 (76%) |
| Women | 6 (60%) | 8 (57%) | 29 (40%) |
| Age, years | 49.1 (31.8 – 57.5) | 34.2 (29.4 – 37.6) | 51.6 (44.9 – 60.2) |
| SARA score | 0.0 (0.0 – 0.0) | 1.0 (0.3 – 1.5) | 10.5 (7.5 – 16.5) |
| Length of expanded CAG allele | n.a. | 69 (65 – 71) | 69 (64 – 71) |
| Time from onset, years | n.a. | -14.6 (-18.1 – -10.7) | 8.3 (4.9 – 13.2) |
| **NfL subgroup (n = 303)** |  |  |  |
| Number of participants | 92 (30%) | 32 (10%) | 179 (59%) |
| Women | 55 (60%) | 18 (56%) | 90 (50%) |
| Age, years | 44.8 (35.7 – 56.6) | 34.2 (26.2 – 39.7) | 51.9 (45.3 – 59.7) |
| SARA score | 0.0 (0.0 – 0.7) | 1.0 (0.5 – 1.6) | 13.0 (8.5 – 20.6) |
| Length of expanded CAG allele | n.a. | 69 (66 – 71) | 69 (66 – 72) |
| Time from onset, years | n.a. | -12.6 (-16.7 – -2.7) | 10.9 (6.4 – 15.9) |
| **MRI subgroup (n = 171)** |  |  |  |
| Number of participants | 45 (26%) | 32 (19%) | 94 (55%) |
| Women | 25 (56%) | 21 (66%) | 38 (40%) |
| Age, years | 45.8 (32.7 – 56.3) | 34.2 (29.5 – 40.8) | 51.8 (45.3 – 58.2) |
| SARA score | 0.0 (0.0 – 0.5] | 1.0 (0.4 – 2.0) | 10.0 (8.0 – 14.5) |
| Length of expanded CAG allele | n.a. | 68. (65 – 71) | 69 (67 – 71) |
| Time from onset, years | n.a. | -12.3 (-17.6 – 2.0) | 8.7 (4.6 – 12.6) |

Data are n, n (%), or median (IQR). NfL=neurofilament light chain. SARA=Scale for the Assessment and Rating of Ataxia.

Data from 856 visits were analysed. Participants had a median number of 2 (IQR 1–3) visits and a median observation time of 1.02 years (0.00 – 2.03). Hundred-and-two SCA3 mutation carriers and 33 healthy controls completed one follow-up visit, 64 mutation carriers and 14 healthy controls two follow-up visits, and 31 mutation carriers and 13 healthy controls three to five follow-up visits. The appendix details the availability of ATXN3, NfL and MRI data at the follow-up visits (appendix p 3).

Baseline results of ATXN3 concentrations, NfL concentrations, and MRI measures are given in appendix p 8. Mutation carriers without ataxia had higher ATXN3 and NfL concentrations, lower medulla oblongata, pons, midbrain, cerebellar white matter (CWM), and superior cerebellar peduncle (SCP) volumes, reduced FA inferior cerebellar peduncle (ICP) and FA SCP, and increased RD ICP values than healthy controls. Because our focus was on early disease stages, we took into account only those MRI measures, which were altered in mutation carriers without ataxia in comparison to healthy controls (appendix p 7). In addition, we included cerebellar grey matter (CGM) volume, to be consistent with the previously published cross-sectional analysis of this cohort.^16^

At baseline, nine of the SCA3 mutation carriers were in the carrier stage, 23 in the biomarker stage, and 236 in the ataxia stage. Twenty-three mutation carriers without ataxia could not be assigned to a disease stage, because NfL concentrations were not available. Within the observation period, two mutation carriers converted from the carrier to the biomarker stage, and seven from the biomarker to the ataxia stage. On the other hand, one mutation carrier assigned to the biomarker stage at baseline was assigned to the carrier stage at the final visit. Further, three mutation carriers, which were scored as ataxic at baseline, were assigned to earlier stages at the final visit: two to the biomarker and one to the carrier stage.

Progression of SARA scores, mutant ATXN3, NfL concentrations, and MRI measures of SCA3 mutation carriers in relation to the time from onset are shown as modelled curves in figure 1. The original data displayed as spaghetti plots are given in the appendix p 9.

**Figure 1:**
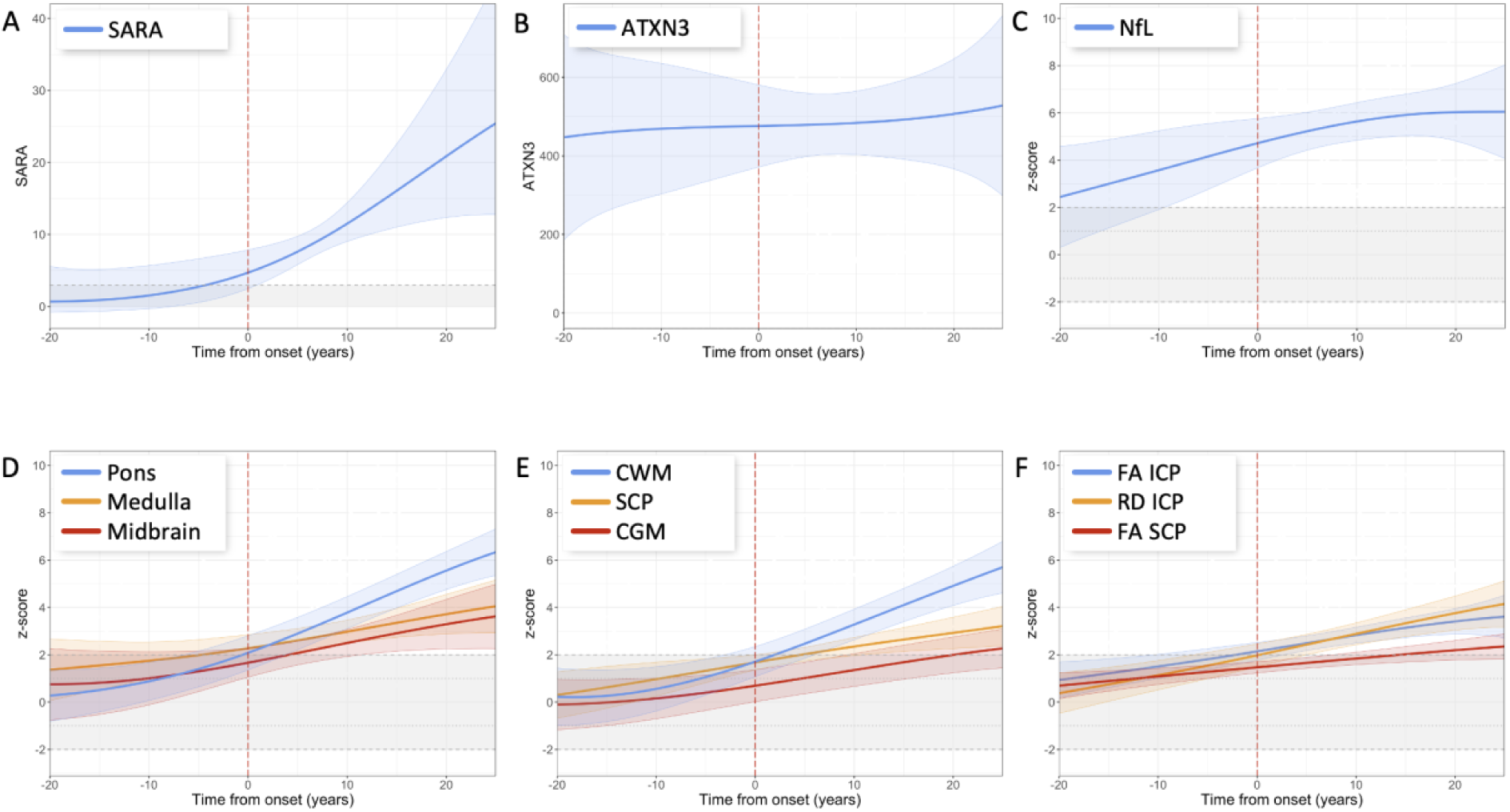
Progression of (A) SARA, (B) ATXN3, (C) NfL, (D) MRI brainstem volumes, (E) MRI cerebellar volumes, and (F) MRI diffusion measures in SCA3. Data were analysed by additive mixed regression models with participant-specific random intercepts on a timescale defined by onset of gait disturbances (vertical dashed line in red) using a cubic P-spline with six B-spline basis functions. The estimated 95% CIs are shown by the shaded areas around the curves. NfL and MRI data were z-transformed in relation to healthy controls. Z-scores of MRI volumes and FA values were inverted for a better visualization. The horizontal ribbon shaded in grey indicates the normal range (±2) of the z-transformed measures (NfL, MRI measures) of healthy controls. For SARA the applied cut-off of 3 is indicated by a dotted horizontal line. CGM=cerebellar grey matter. CWM=cerebellar white matter. FA ICP=fractional anisotropy of the inferior cerebellar peduncle. FA SCP=fractional anisotropy of the superior cerebellar peduncle. NfL=neurofilament light chain. RD ICP=radial diffusivity of the inferior cerebellar peduncle. SCP= superior cerebellar peduncle.

SARA progression had a sigmoidal shape (figure 1A). Scores crossed the cut-off of 3, which defines the onset of the ataxia stage, 4.2 years [95% CI n.d. – 0.9] before the reported or estimated onset of gait disturbances (table 2). At the time of onset of gait disturbances, the SARA score was 4.7 [2.5 – 7.9]. Mutant ATXN3 concentrations were constant throughout the entire disease course without major changes over time so that the onset of abnormality could not be determined (figure 1B). NfL concentrations increased throughout the disease course, but the increase slowed down after onset (figure 1C). NfL values became abnormal more than 20 years (-21.5 years [n.d. – -9.5]) before onset (table 2). All analysed MRI volumes and diffusion measures worsened over time, albeit at different rates and with different slopes (figure 1D-F). Medulla oblongata volume (-4.7 years [n.d. – +3.7]), FA ICP (-2.1 years [-10.3 – 2.7]), and pons volume (-0.6 years [-6.4 – 3.8]) became abnormal before onset. The remaining MRI measures became abnormal 0.3 to 14.4 years after onset in the following temporal order: RD ICP, CWM volume, midbrain volume, SCP volume, and FA SCP. CGM value did not decrease more than 2 SD below the mean of healthy controls throughout the entire disease course (table 2). Compared to our previous cross-sectional analysis, in which we determined the upper 95% CI limits of NfL, pons volume, and CWM volume, the onset of abnormality of these measures was 3.8 to 6.5 years later. To clarify the reason for this difference, we calculated the upper 95% CI limits of NfL, pons volume, and CWM volume based alone on the baseline values of the present dataset. These values differed only by 1.1 to 3.2 years from the previously determined values (appendix p 12).

**Table 2:**
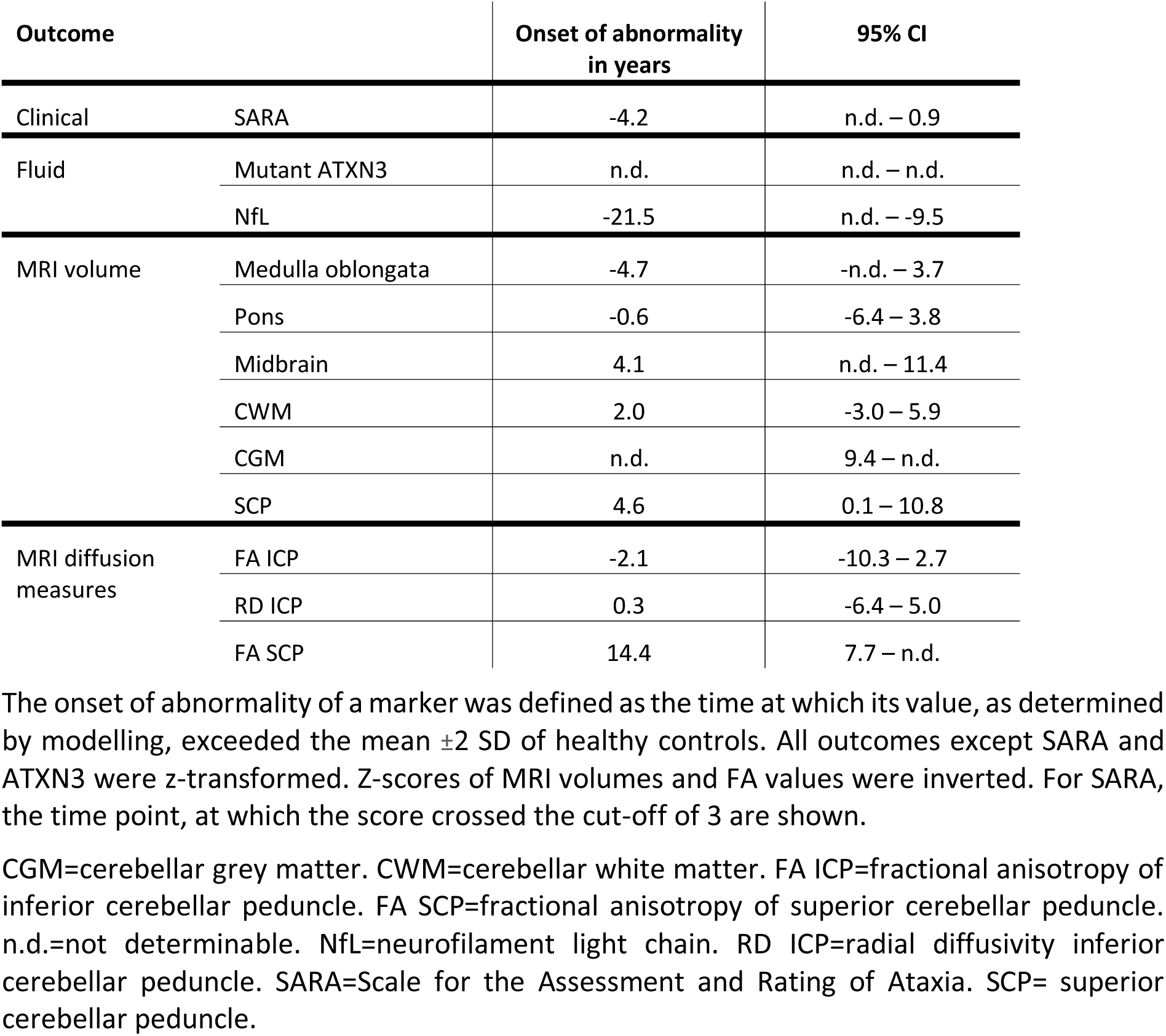
Onset of abnormality of SARA, ATXN3, NfL, and MRI measures in SCA3.

| Outcome |  | Onset of abnormality<br>in years | 95% CI |
| --- | --- | --- | --- |
| Clinical | SARA | -4.2 | n.d. – 0.9 |
| Fluid | Mutant ATXN3 | n.d. | n.d. – n.d. |
|  | NfL | -21.5 | n.d. – -9.5 |
| MRI volume | Medulla oblongata | -4.7 | -n.d. – 3.7 |
|  | Pons | -0.6 | -6.4 – 3.8 |
|  | Midbrain | 4.1 | n.d. – 11.4 |
|  | CWM | 2.0 | -3.0 – 5.9 |
|  | CGM | n.d. | 9.4 – n.d. |
|  | SCP | 4.6 | 0.1 – 10.8 |
| MRI diffusion<br>measures | FA ICP | -2.1 | -10.3 – 2.7 |
|  | RD ICP | 0.3 | -6.4 – 5.0 |
|  | FA SCP | 14.4 | 7.7 – n.d. |

The onset of abnormality of a marker was defined as the time at which its value, as determined by modelling, exceeded the mean $\pm 2$ SD of healthy controls. All outcomes except SARA and ATXN3 were z-transformed. Z-scores of MRI volumes and FA values were inverted. For SARA, the time point, at which the score crossed the cut-off of 3 are shown.

To determine the responsiveness of the studied measures, we calculated the SCSs. For the entire disease, all measures except ATXN3 had SCSs larger than 0. The most sensitive measure was pons volume with an SCS of 1.35 [95% CI 1.11 – 1.78]. Further stage-specific analyses showed that the SCSs of the various outcome measures depended on the disease stage. In the carrier stage, the SCSs of SCP volume (0.62 [ 0.04 – 1.05]) and FA SCP (0.45 [ 0.17 – 0.92]) were larger than 0, whereas the SCSs of SARA and all other analysed biological markers did not differ from 0. In the biomarker stage, SCSs of SARA, NfL, all MRI volumes except midbrain, and RD ICP were larger than 0. In this stage, pons volume had the highest SCS of all outcome measures (1.41 [ 0.64 – 3.29]), followed by SCP (0.81 [ 0.34 – 1.64]) and CWM volume (0.78 [ 0.11 – 1.73]). Pons volume also had the highest SCS in the ataxia stage (1.71 [ 1.32 – 2.45]). It markedly exceeded the SCS of SARA (0.69 [ 0.60 – 0.80]). In the ataxia stage, the SCSs of NfL and FA SCP did not differ from 0, whereas all other biological markers had SCSs larger than 0 (table 3).

**Table 3:** Stage-specific sensitivity to change (SCS) of SARA, ATXN3, NfL, and MRI measures in SCA3.

| Outcome measure |  | Entire disease | Carrier stage | Biomarker stage | Ataxia stage |
| --- | --- | --- | --- | --- | --- |
| Clinical | SARA | 0.99 | 0.07 | 0.37 | 0.69 |
|  |  | (0.88 – 1.11) | (-0.25 – 0.52) | (0.19 – 0.59) | (0.60 – 0.80) |
| Fluid | Mutant ATXN3 | 0.04 | 0.09 | -0.11 | 0.10 |
|  |  | (-0.11 – 0.20) | (-0.78 – 7.61) | (-1.55 – 0.72) | (-0.01 – 0.21) |
|  | NfL | 0.21 | 0.51 | 0.63 | 0.08 |
|  |  | (0.14 – 0.30) | (-0.11 – 1.28) | (0.37 – 0.99) | (-0.03 – 0.19) |
| MRI volume | Medulla oblongata | 0.48 | 0.20 | 0.52 | 0.30 |
|  |  | (0.33 – 0.73) | (-0.56 – 0.60) | (0.04 – 1.20) | (0.13 – 0.75) |
|  | Pons | 1.35 | 0.19 | 1.41 | 1.71 |
|  |  | (1.11 – 1.78) | (-0.11 – 0.50) | (0.64 – 3.29) | (1.32 – 2.45) |
|  | Midbrain | 0.49 | -0.18 | 0.31 | 0.40 |
|  |  | (0.34 – 0.80) | (-0.59 – 0.12) | (-0.17 – 0.89) | (0.16 – 1.00) |
|  | CWM | 1.01 | - 0.03 | 0.78 | 1.10 |
|  |  | (0.85 – 1.23) | (-0.54 – 0.22) | (0.11 – 1.73) | (0.84 – 1.48) |
|  | CGM | 0.64 | 0.00 | 0.48 | 0.72 |
|  |  | (0.50 – 0.80) | (-0.25 – 0.49) | (0.19 – 0.79) | (0.54 – 0.93) |
|  | SCP | 0.69 | 0.62 | 0.81 | 0.30 |
|  |  | (0.52 – 0.94) | (0.04 – 1.05) | (0.34 – 1.64) | (0.14 – 0.53) |
| MRI diffusion measures | FA ICP | 0.56 | 0.11 | 0.24 | 0.17 |
|  |  | (0.44 – 0.67) | (-0.22 – 0.40) | (-0.03 – 0.58) | (0.03 – 0.32) |
|  | RD ICP | 0.56 | 0.17 | 0.51 | 0.24 |
|  |  | (0.44 – 0.71) | (-0.08 – 0.70) | (0.17 – 1.02) | (0.09 – 0.40) |
|  | FA SCP | 0.38 | 0.45 | 0.26 | 0.04 |
|  |  | (0.22 – 0.58) | (0.17 – 0.92) | (-0.06 – 0.54) | (-0.11 – 0.23) |

Data are the estimated slope of progression (95% CI).

To identify factors that predicted SARA progression we applied univariable and multivariable modelling. In the univariable analysis of the entire disease, lower age, larger CAG repeat length, and lower volumes of medulla oblongata, midbrain, CGM, and SCP were associated with faster SARA progression. In the carrier stage, lower age, female sex, higher ATXN3 levels, larger CAG repeat length, and lower medulla oblongata and CWM volumes were predictors, in the ataxia stage, lower age, larger CAG repeat length, and lower medulla oblongata, midbrain and CGM volumes. In the biomarker stage, we did not find significant predictors (appendix pp 10-11). The multivariable analysis of the entire disease course selected lower age (p=0.0459) and lower medulla oblongata volume (p<0.0001) as predictors of SARA progression (figure 2).

**Figure 2.**
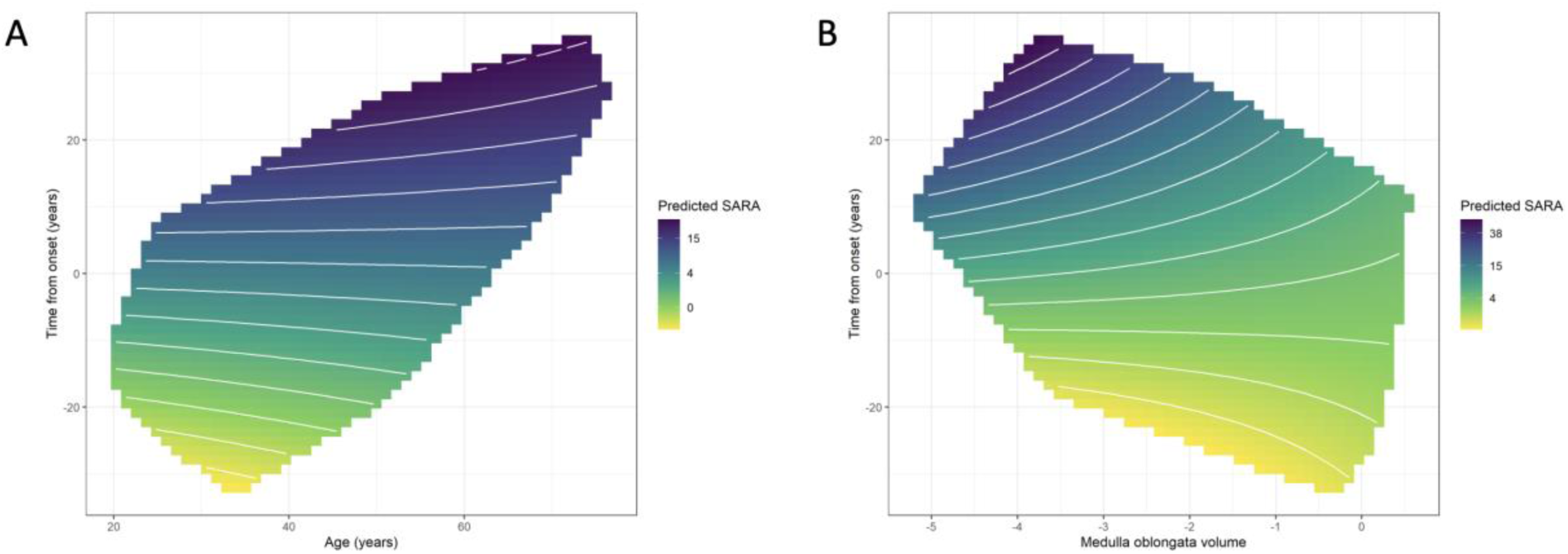
Partial dependence plots of the multivariable model for SARA progression. Predicted values of SARA as a function of the time from onset and age (A) and the time from onset and medulla oblongata volume given as z-score (B). In both panels, the value of the other predictor was set to the observed mean (medulla oblongata volume: -2.6; age: 46.4 years), respectively. The closer the white lines, which represent identical SARA values, are together, the faster is the predicted progression.

## Discussion

This longitudinal study determined the sequence and extent of plasma mutant ATXN3, plasma NfL and MRI outcome measure changes along the SCA3 disease course in participants of the ESMI cohort. We analysed the data in the framework of the recently proposed SCA3 staging model that distinguishes an asymptomatic carrier stage, a biomarker stage and the final ataxia stage.^16^

The present analysis showed that pre-ataxic SCA3 mutation carriers on average entered the biomarker stage 21.5 years before clinical onset with an upper margin of the 95% CI of 9.5 years before onset. The moderate difference to the previously reported time of 13.3 years is most likely due to the fact that we now analysed longitudinal data, while the previous analysis was based on cross-sectional data.^16^ The ataxia stage started 4.2 years before the onset, defined by the estimated or reported onset of gait abnormalities. In the RISCA study, the observed conversion of SCA mutation carriers to ataxia also occured before the estimated onset.^22^ These data provide convergent evidence that the clinically determined ataxia onset precedes the self-perceived onset, which is generally equated with the clinical onset. The number of observed transitions between stages was low. This reflects the relatively slow progression of SCA3. The low number of stage transitions is also due to individual fluctuations of outcome measures. Indeed, video-based home recordings of SARA revealed short-term fluctuations of several score points.^23^

Mutant ATXN3 concentrations were almost constant across the entire disease course and did not show relevant dynamics. Correspondingly, ATXN3 provides no information about the progression of SCA3. However, ATXN3 has potential as a target engagement marker in gene silencing trials.^3,4^ NfL levels became abnormal earlier than any of the analysed MRI measures. NfL slowly increased and stayed at elevated levels throughout the disease course. As NfL reflects the rate of neurodegeneration rather than disease severity, constantly increased NfL levels indicate ongoing disease progression. NfL might therefore be studied as a treatment response marker for SCA3.^5,8^

The earliest MRI abnormalities were volume loss of the medulla oblongata and reduced FA ICP, followed by volume loss of the pons and increased RD ICP, while cerebellar measures became abnormal only in the later course. The ICP contains the dorsal spinocerebellar tract and fibre tracts connecting the medulla oblongata with the cerebellum. Together with previous reports of impaired microstructural integrity of the ICP in pre-ataxic SCA3 mutation carriers,^15,24^ these findings suggest a pathological process that originates in the spinal cord and lower brainstem and further ascends to the cerebellum. They further indicate early white matter pathology in SCA3. This is in line with the observation of impaired oligodendrocyte maturation in two animal models of SCA3.^25,26^

To assess the responsiveness of SARA and the analysed biological markers, we determined the SCSs for each of them. This analysis revealed stage-dependence of SCSs. In both, biomarker and ataxia stage as well as across the entire disease course, pons volume had the highest SCS of all analysed measures. The superior responsiveness of the MRI volume measures compared to SARA is in line with previous studies in small cohorts of ataxic SCA3 individuals.^11,12^ Our results agree with those of a prospective MRI study of 24 SCA3 mutation carriers over 6 months in that MRI measures were more sensitive to change than SARA and that pons volume had the highest responsiveness of the studied MRI measures.^14^ The responsiveness of NfL was low across the entire disease course. This is in agreement with a previous longitudinal study in 19 SCA3 patients that did not find a NfL increase over two years.^9^

This study not only investigated the influence of biological factors, such as age, sex, and CAG repeat length, on disease progression in SCA3, but also that of fluid and MRI markers. Some, but not all previous studies in SCA3 reported an association between the length of the expanded CAG repeat and faster progression of ataxia severity.^27–30^ In addition, greater CAG repeat length was reported to be a risk factor for the conversion of pre-ataxic SCA3 individuals to manifest ataxia.^22^ In the univariable analysis of the present data, greater CAG repeat length and lower age were associated with faster SARA progression. In the multivariable analysis, lower age was one of two selected factors. As CAG repeat length and age of onset are inversely correlated in SCA3,^1^ the opposing effects of CAG repeat length and age suggest a biological effect of the expansion size on the dynamics of disease progression. Of all biological markers investigated, only MRI volume measures were identified as predictors of progression. Among them, lower medulla oblongata volume had the most consistent effect.

A main limitation of this study is the small number of observed stage transitions. We were therefore not in the position to identify predictors of progression, as indicated by transition to more advanced disease stages. Another limitation is that the study was conducted mainly with European participants. It is thus unclear, whether the results can be generalized to SCA3 mutation carriers from other world regions.

In conclusion, our study provides quantitative information on the progression of biological markers in SCA3 mutation carriers before and after onset of ataxia, and allowed the identification of predictors for clinical progression. Our data are useful for the design of future clinical trials. Of particular importance is the finding that pons volume was more sensitive to change than any other outcome. This characterizes pons volume as a useful marker to monitor progression in clinical trials.

## Data Availability

All data produced in the present study are available upon reasonable request to the authors.

## Acknowledgements

Several authors of this publication are members of the European Reference Network for Rare Neurological Diseases. JF was funded within the Advanced Clinician Scientist Programme (ACCENT, funding code 01EO2107, by the German Federal Ministry of Education and Research (BMBF) and as a PI of the iBehave Network, sponsored by the Ministry of Culture and Science of the State of North Rhine-Westphalia. MMS and LA were funded by the European Regional Development Fund (ERDF), through the Centro 2020 Regional Operational Programme under project CENTRO-01-0145-FEDER-181240 and through the COMPETE 2020 - Operational Programme for Competitiveness and Internationalisation and Portuguese national funds via FCT – Fundação para a Ciência e a Tecnologia, under projects 2022.06118.PTDC, UIDB/04539/2020, UIDP/04539/2020 and LA/P/0058/2020, and European funds (GeneT GA 101059981) and by National Ataxia Foundation (NAF). MGE received research support from the German Ministry of Education and Research (BMBF) within the European Joint Program for Rare Diseases (EJP-RD) 2021 Transnational Call for Rare Disease Research Projects (funding number 01GM2110), from the National Ataxia Foundation (NAF), and from Ataxia UK, all unrelated to the present manuscript. MR is supported by Fundação para a Ciência e Tecnologia (FCT; CEECIND/03018/2018/CP1556/CT0009). In the Netherlands, this work was supported by ZonMw (grant number 733051066). The Center for Magnetic Resonance Research is supported by the National Institute of Biomedical Imaging and Bioengineering (NIBIB) grant P41 EB027061, the Institutional Center Cores for Advanced Neuroimaging award P30 NS076408 and S10 OD017974 grant. ER was supported in part by the National Ataxia, Great Oaks, Once Upon A Time, and MINDlink Foundations. PS was funded by FCT under the grant SFRH/BD/148451/2019. DT received funding from the DFG, EU and Bernd Fink Foundation unrelated to the study. FE received funding from the DFG in the framework of the DFG Clinician Scientist Programme UMEA, FU 356/12-2. HZ is a Wallenberg Scholar supported by grants from the Swedish Research Council (#2018-02532), the European Research Council (#681712), Swedish State Support for Clinical Research (#ALFGBG-720931), the Alzheimer Drug Discovery Foundation (ADDF), USA (#201809-2016862), and the UK Dementia Research Institute at UCL. PG is supported by the National Institute for Health Research University College London Hospitals Biomedical Research Centre UCLH. PG receives also support from the North Thames CRN. PG and HGM, work at University College London Hospitals/ University College London, which receives a proportion of funding from the Department of Health’s National Institute for Health Research Biomedical Research Centre’s funding scheme. PG received funding from CureSCA3 in support of HGM work. LS received funding by the Deutsche Forschungsgemeinschaft (DFG, grants SCHO754/6-2 and SCA754/8-1), the German Ministry of Education and Research (BMBF, grants 01GM2209F and 01GM2210A) and the German Ministry of Health (BMG, grant ZMVI1-2520DAT94E) and the European Union (grant 947588), all unrelated to this project. MS is supported by the European Union, project European Rare Disease Research Alliance (ERDERA), GA n°101156595, funded under call HORIZON-HLTH-2023-DISEASE-07.

## Appendix

**Appendix Figure 1:**
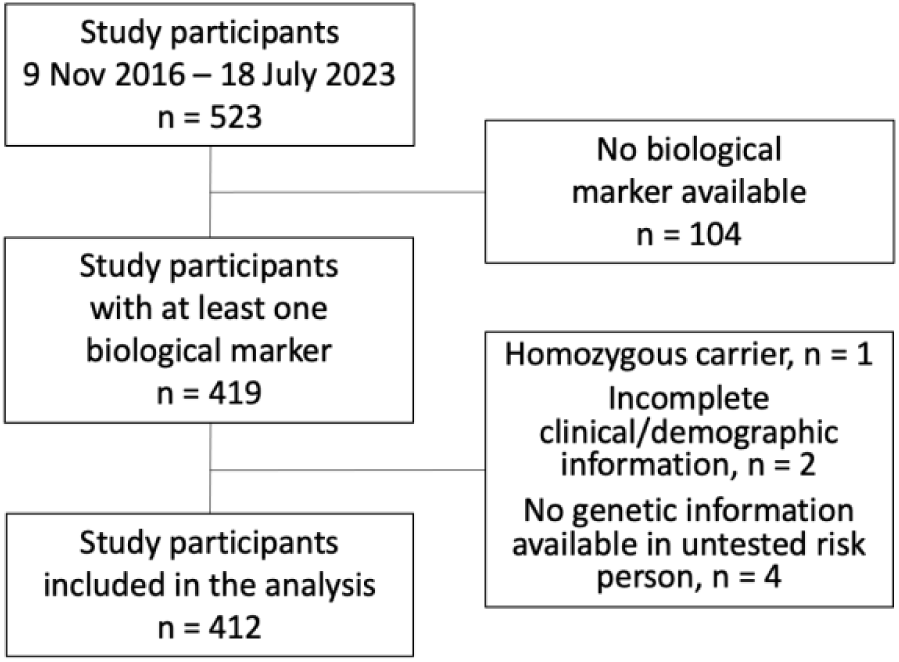
Flow chart of study participants

In total, 523 participants were enrolled in the European Spinocerebellar ataxia type 3/Machado-Joseph Disease Initiative (ESMI) from Nov 9,2016 to July 18, 2023. For 104 participants no fluid or imaging biomarker was available. From the remaining 419 participants, 412 were included in the present analysis.

**Appendix Table 1:** Exclusion of visit-specific individual data of biological markers.

| Biological marker | Reason | n |
| --- | --- | --- |
| ATXN3 | Values out of detection range | 5 |
|  | Not enough biosample volume left for the repetition of measurement, that was necessary due to a failure in the first measurement | 9 |
|  | Outlier value (ATXN3: 2277 pg/mL*dilution factor) | 1 |
| NfL | Missing visit date information | 3 |
|  | Outlier value (NfL level of >50 pg/mL in healthy controls (56.99 and 335.10) and >150 pg/mL in mutation carriers (168.43, 293.02, 360.88)) | 5 |
| MRI | Date difference between MRI and clinical assessment > 60 days | 10 |
|  | Failure in quality control | 3 |
|  | Failure in segmentation pipeline | 2 |
|  | Outlier value (Pons: 8.7 ‰ eTIV) | 1 |
| Total |  | 39 |

In total 39 individual biological marker measurements had to be excluded from the analysis.

**Appendix Table 2:** Data availability.

|  | Healthy controls | SCA3 mutation carriers with<br>SARA < 3 | SCA3 mutation carriers with<br>SARA ≥ 3 |
| --- | --- | --- | --- |
| Baseline | 10 ATXN3<br>92 NfL<br>45 MRI | 14 ATXN3<br>32 NfL<br>32 MRI | 73 ATXN3<br>179 NfL<br>94 MRI |
| 1 follow-up visit | 9 ATXN3<br>42 NfL<br>27 MRI | 12 ATXN3<br>22 NfL<br>20 MRI | 70 ATXN3<br>132 NfL<br>45 MRI |
| 2 follow-up visits | 4 ATXN3<br>9 NfL<br>19 MRI | 1 ATXN3<br>9 NfL<br>11 MRI | 21 ATXN3<br>54 NfL<br>26 MRI |
| ≥3 follow-up visits | 0 ATXN3<br>0 NfL<br>12 MRI | 0 ATXN3<br>0 NfL<br>6 MRI | 0 ATXN3<br>6 NfL<br>18 MRI |

### Appendix Methods: MRI protocol and analysis pipelines

All scans were acquired on Siemens 3T scanners (Siemens Medical Systems, Erlangen, Germany). T1-weighted MRIs were acquired using a magnetization prepared rapid gradient-echo sequence (MPRAGE, TR = 2500 ms, TE = 4.37 ms, TI = 1100 ms, flip angle = 7 deg, FOV 256 mm x 256 mm, 192 slices with a voxel size of 1 mm isotropic). Diffusion weighted images were acquired using a twice-refocused, single-shot, echo planar imaging sequence (TR = 12100 ms, TE = 88 ms, FOV = 240 mm x 240 mm, 72 axial slices) with a voxel size of 2 mm isotropic, two diffusion-weighted imagines with b-value of 700/mm2 and 1000s/mm^2^ were acquired at 30 diffusion-encoding directions, respectively).

Brain volumes and the estimated intracranial volume were calculated using FastSurfer^1^, brainstem volumes were calculated using FreeSurfer^2^ and cerebellar volumes were calculated using CerebNet^3^. All volumes have been normalized by the estimated intracranial volume (eTIV) as follows: volume_norm_ = volume / eTIV. For hemispherical volumes, the average of both hemispheres was calculated. Only normalized volumes were considered for the statistical analysis.

The DWI data were pre-processed using MRtrix3 which involved denoising and correction for eddy currents, susceptibility-induced distortions, and bias field.^4–6^ Based on the diffusion tensor, we calculated the following metrics: fractional anisotropy (FA), medial diffusivity (MD), axial (AD) and radial diffusivity (RD) to study white matter integrity. Diffusion data harmonization was done with neuroCombat.^7,8^ Mean values of the diffusion metrics were calculated for 14 white matter tracts.

A full list of all considered volumes and tracts is given in appendix table 3.

**Appendix Table 3:**
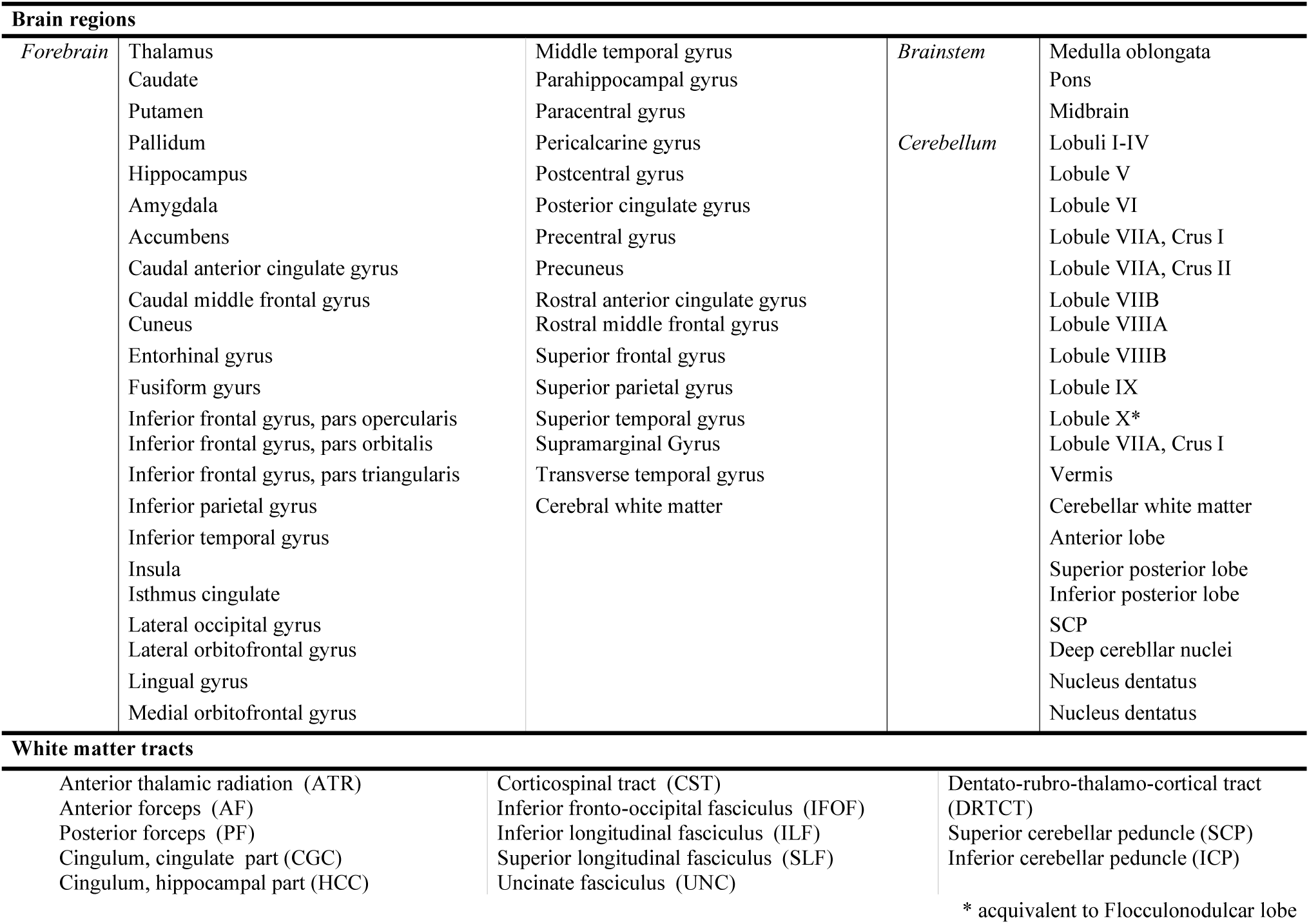
Anatomical structures analysed by MRI.

### Appendix Methods: Statistical analysis for selection of biological markers

The selection of studied imaging biomarker was based on the statistical group comparison of the cross-sectional MRI at baseline. Statistical analyses were performed with R version 4.3.1. The analysis aims to assess the effect of group membership (ataxic SCA3 mutation carriers, pre-ataxic mutation carriers, and healthy controls) on the dependent variable (each brain volume and diffusion volume), adjusting for the covariates age and sex. Separate linear models are applied to estimate the effects of group differences while controlling for these covariates age and sex for each brain volume and MRI diffusion measure. Post-hoc pairwise comparisons between groups are conducted using Tukey contrasts (with Rpackage multcomp version1.4-20) to identify significant differences (p<=0.05) while adjusting p-values for multiple comparisons.

To address multiple testing, two complementary approaches are applied; this two-step correction ensures both a global assessment of group differences (via Tukey contrasts) and a more focused analysis of pre-ataxic versus healthy controls (via Benjamini-Hochberg adjustment). Combining these approaches provides a robust framework for identifying both broad and specific patterns of significance while minimizing the risk of false-positive results.

The assumptions underlying the statistical methods, including linearity, homoscedasticity, normality of residuals, and independence of observations for the linear regression model, as well as homogeneity of variances for Tukey’s HSD, were tested and confirmed to be met, ensuring the validity of the analysis. These checks provide confidence that the results are robust and interpretable.

We report the statistics for the significant single contrast pre-ataxic SCA3 mutation carrier with a SARA < 3 versus healthy controls (and p<=0.01). Estimate, standard errors (SE), t-values and Benjamini-Hochberg corrected p-values are given in appendix table 4.

**Appendix Table 4:**
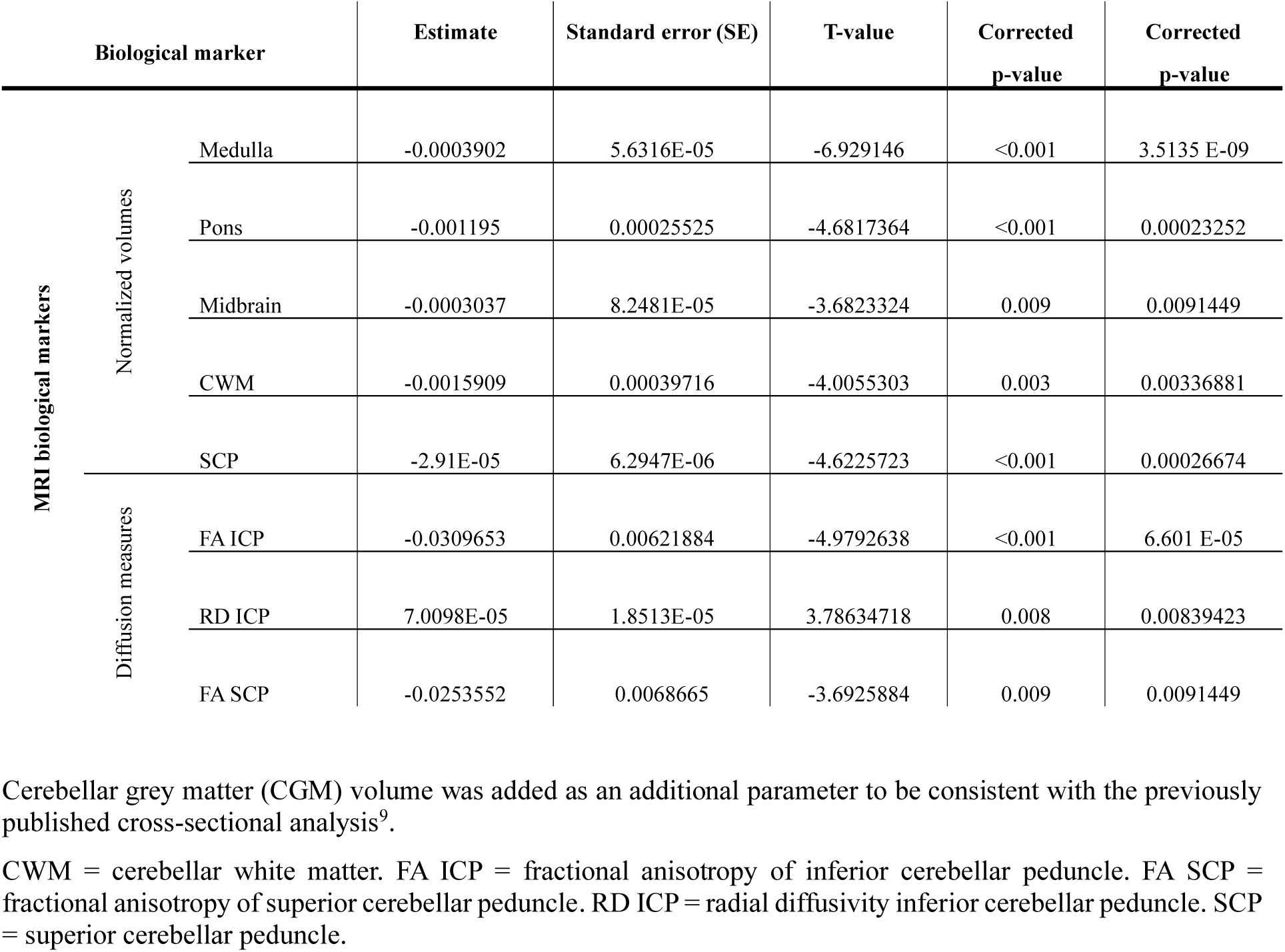
Estimates, standard errors, T-values and corrected p-values of the selected MRI markers.

**Appendix Table 5:**
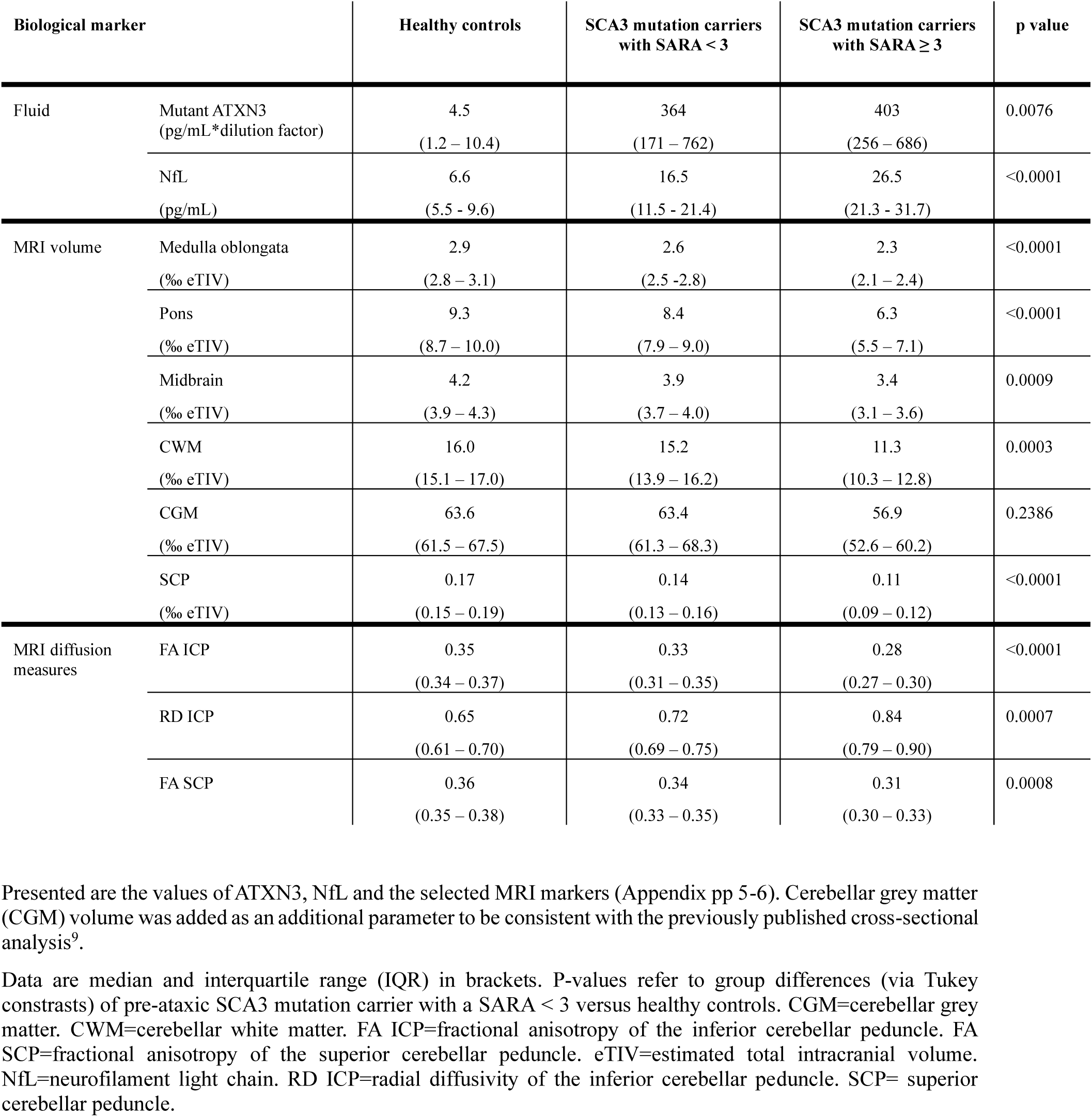
Baseline values of ATXN3, NfL, and selected MRI measures.

**Appendix Figure 2:**
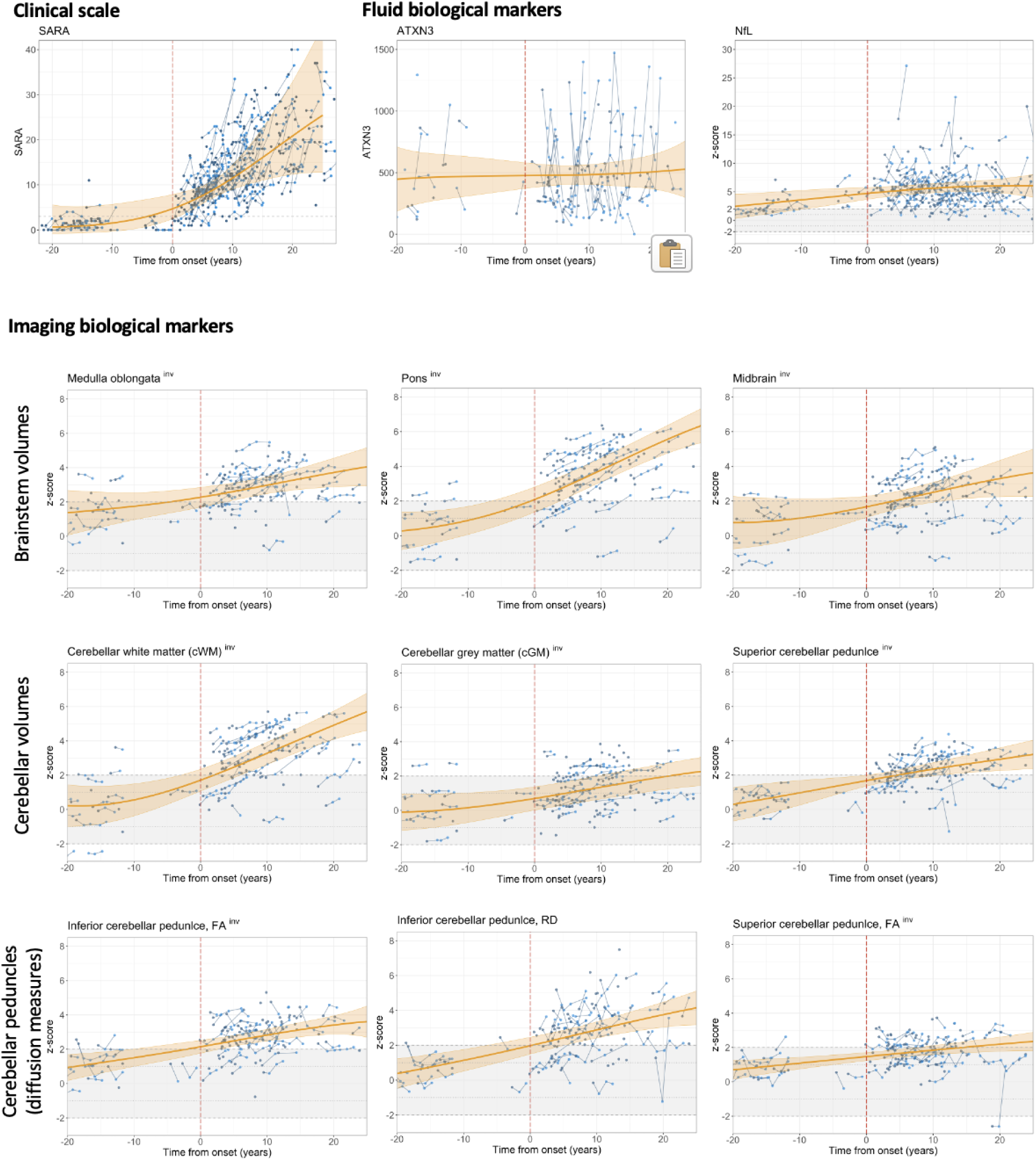
Individual trajectories of SARA and biological markers in SCA3 Data are shown as individual trajectories of SARA, fluid biological markers, and MRI markers in SCA3 on a timescale defined by onset of gait disturbances. For illustration, the modelled curves shown in figure 1 of the main text are superimposed. NfL and MRI data were z-transformed in relation to healthy controls. Z-scores of MRI volumes and FA values were inverted. The normal distribution in healthy controls defined by the range of z-scores between ± 2 is indicated by the horizontal ribbon shaded in grey. For the SARA sum score the cut-off value of 3^10,11^ is indicated by a dotted horizontal line. CGM=cerebellar grey matter. CWM=cerebellar white matter. FA ICP=fractional anisotropy of inferior cerebellar peduncle. FA SCP=fractional anisotropy of superior cerebellar peduncle. NfL=neurofilament light chain. RD ICP=radial diffusivity inferior cerebellar peduncle. SARA=Scale for the Assessment and Rating of Ataxia. SCP= superior cerebellar peduncle.

**Appendix Table 6:** Baseline predictors of SARA progression in SCA3.

| Variable | Entire disease |  | Carrier stage |  | Biomarker stage |  | Ataxia stage |  |
| --- | --- | --- | --- | --- | --- | --- | --- | --- |
|  | Slope change [95% CI] | p value | Slope change [95% CI] | p value | Slope change [95% CI] | p value | Slope change [95% CI] | p value |
| Age | -0.000<br>[-0.0013 – 0.0000] | 0.0394 | -0.0091<br>[-0.0166 – -0.0021] | 0.0245 | -0.0004<br>[-0.0030 – 0.0021] | 0.7585 | -0.0013<br>[-0.0023 – -0.0004] | 0.0073 |
| Sex | -0.0161<br>[-0.0326 – 0.0004] | 0.0570 | -0.1183<br>[-0.2014 – -0.0326] | 0.0107 | -0.0414<br>[-0.0885 – 0.0056] | 0.1060 | -0.0064<br>[-0.0298 – 0.0169] | 0.5919 |
| CAG repeat length, longer allele | 0.0060<br>[0.0042 – 0.0078] | 0.0000 | 0.0091<br>[0.0017 – 0.0168] | 0.0277 | 0.0011<br>[-0.0040 – 0.0062] | 0.6901 | 0.0059<br>[0.0034 – 0.0084] | 0.0000 |
| Mutant ATXN3 | 0.0000<br>[0.0000 – 0.0001] | 0.0899 | 0.0006<br>[0.0002 – 0.0011] | 0.0204 | 0.0001<br>[-0.0002 – 0.0005] | 0.5950 | 0.0001<br>[0.000 – 0.0001] | 0.1554 |
| NfL | 0.0005<br>[-0.0029 – 0.0039] | 0.7700 | -0.0825<br>[-0.4661 – 0.3297] | 0.6767 | 0.0032<br>[-0.0171 – 0.0232] | 0.7716 | -0.0016<br>[-0.0056 – 0.0023] | 0.4146 |
| Medulla oblongata volume | -0.0180<br>[-0.0268 – -0.0092] | 0.0001 | -0.0522<br>[-0.0965 – -0.0108] | 0.0415 | -0.0087<br>[-0.0378 – 0.0204] | 0.5902 | -0.0332<br>[-0.0511 – -0.0145] | 0.0001 |
| Pons volume | -0.0050<br>[-0.0110 – 0.0010] | 0.1017 | 0.0240<br>[-0.0405 – 0.0791] | 0.4406 | -0.0029<br>[-0.0261 – 0.0203] | 0.8233 | -0.0038<br>[-0.0114 – 0.0040] | 0.3314 |
| Midbrain volume | -0.0122<br>[-0.0203 – -0.0041] | 0.0037 | -0.0131<br>[-0.0724 – 0.0386] | 0.6644 | -0.0060<br>[-0.0357 – 0.0237] | 0.7192 | -0.0113<br>[-0.0213 – -0.0013] | 0.0227 |
| CWM volume | -0.0058<br>[-0.0117 – 0.0001] | 0.0561 | 0.0361<br>[0.0014 – 0.0677] | 0.0450 | -0.0013<br>[-0.0171 – 0.0145] | 0.8805 | -0.0069<br>[-0.0171 – 0.0035] | 0.1781 |
| CGM volume | -0.0127<br>[-0.0218 – -0.0037] | 0.0059 | 0.0289<br>[-0.0487 – 0.1061] | 0.5039 | -0.0060<br>[-0.0260 – 0.0142] | 0.5872 | -0.0268<br>[-0.0463 – -0.0066] | 0.0027 |
| SCP volume | -0.0138<br>[-0.0234 – -0.0042] | 0.0053 | -0.0691<br>[-0.1865 – 0.0491] | 0.2975 | -0.0099<br>[-0.0366 – 0.0167] | 0.5082 | -0.0050<br>[-0.0264 – 0.0166] | 0.6464 |
| FA ICP | -0.0085<br>[-0.0180 – 0.0010] | 0.0835 | 0.0164<br>[-0.0594 – 0.0862] | 0.6821 | -0.0349<br>[-0.0858 – 0.0171] | 0.2294 | 0.0173<br>[-0.0056 – 0.0394] | 0.1267 |
| RD ICP | 0.0059<br>[-0.0026 – 0.0144] | 0.1721 | -0.0006<br>[-0.1108 – 0.1225] | 0.9927 | 0.0053<br>[-0.0273 – 0.0380] | 0.7709 | -0.0012<br>[-0.0203 – 0.0179] | 0.9051 |
| FA SCP | -0.0078<br>[-0.0211 – 0.0055] | 0.2474 | -0.0621<br>[-0.1448 – 0.0196] | 0.1922 | -0.0668<br>[-0.1603 – 0.0553] | 0.1839 | 0.0157<br>[-0.0074 – 0.0384] | 0.1813 |

Shown are the results of the univariable analysis.

**Appendix Table 7:** Comparison of the onset of abnormality data of the present analysis with those of our previous cross-sectional analysis. ^9^

| Parameter | Onset of abnormality in years (upper limit of 95% CI) |  |  |
| --- | --- | --- | --- |
|  | Present analysis<br>All data | Present analysis<br>Only (cross-sectional) baseline data | Previous analysis cross-sectional<br>data<sup>9</sup> |
| NfL | -9.5 | -12.2 | -13.3 |
| Pons volume | 3.8 | 0.1 | -2.2 |
| CWM volume | 5.9 | 2.6 | -0.6 |

The onset of abnormality of a biological marker was defined as the timepoint at which its value, as determined by modelling, exceeded the mean $\pm 2$ SD of healthy controls. To allow direct comparison with our previous, cross-sectional study<sup>9</sup>, we report the intersection with the upper limits of 95% CIs for the present analysis (left data column), the present analysis based alone on the cross-sectional baseline data (middle data column), and the results of our previous analysis (right data column), which was based on a largely overlapping, cross-sectional analysis of the ESMI cohort with a data freeze at Jan 31, 2022.
CWM=cerebellar white matter. NfL=neurofilament light chain.

## References

1 Paulson H. Machado-Joseph disease/spinocerebellar ataxia type 3. Handb. Clin. Neurol. 2012; 103: 437–49.

2 Klockgether T, Ashizawa T, Brais B, et al. Paving the Way Toward Meaningful Trials in Ataxias: An Ataxia Global Initiative Perspective. Mov Disord 2022; 37: 1125–30. 10.1002/mds.29032.

3 Hübener-Schmid J, Kuhlbrodt K, Peladan J, et al. Polyglutamine-Expanded Ataxin-3: A Target Engagement Marker for Spinocerebellar Ataxia Type 3 in Peripheral Blood. Mov Disord 2021; 36: 2675–81. 10.1002/mds.28749.

4 Prudencio M, Garcia-Moreno H, Jansen-West KR, et al. Toward allele-specific targeting therapy and pharmacodynamic marker for spinocerebellar ataxia type 3. Sci. Transl. Med. 2020; 12. 10.1126/scitranslmed.abb7086.

5 Gaetani L, Blennow K, Calabresi P, Di Filippo M, Parnetti L, Zetterberg H. Neurofilament light chain as a biomarker in neurological disorders. J. Neurol. Neurosurg. Psychiatry 2019; 90: 870–81. 10.1136/jnnp-2018-320106.

6 Du Tezenas Montcel S, Petit E, Olubajo T, et al. Baseline Clinical and Blood Biomarkers in Patients With Preataxic and Early-Stage Disease Spinocerebellar Ataxia 1 and 3. Neurology 2023; 100: e1836–e1848. 10.1212/WNL.0000000000207088.

7 Garcia-Moreno H, Prudencio M, Thomas-Black G, et al. Tau and neurofilament light-chain as fluid biomarkers in spinocerebellar ataxia type 3. Eur J Neurol 2022; 29: 2439–52. 10.1111/ene.15373.

8 Wilke C, Haas E, Reetz K, et al. Neurofilaments in spinocerebellar ataxia type 3: blood biomarkers at the preataxic and ataxic stage in humans and mice. EMBO Mol. Med. 2020; 12: e11803. 10.15252/emmm.201911803.

9 Coarelli G, Darios F, Petit E, et al. Plasma neurofilament light chain predicts cerebellar atrophy and clinical progression in spinocerebellar ataxia. Neurobiol. Dis. 2021; 153: 105311. 10.1016/j.nbd.2021.105311.

10 Li Q-F, Dong Y, Yang L, et al. Neurofilament light chain is a promising serum biomarker in spinocerebellar ataxia type 3. Mol. Neurodegener. 2019; 14: 39. 10.1186/s13024-019-0338-0.

11 Adanyeguh IM, Perlbarg V, Henry PG, et al. Autosomal dominant cerebellar ataxias: Imaging biomarkers with high effect sizes. Neuroimage. Clin. 2018; 19: 858–67.

12 Reetz K, Costa AS, Mirzazade S, et al. Genotype-specific patterns of atrophy progression are more sensitive than clinical decline in SCA1, SCA3 and SCA6. Brain 2013; 136: 905–17.

13 Piccinin CC, Rezende TJR, Paiva JLR de, et al. A 5-Year Longitudinal Clinical and Magnetic Resonance Imaging Study in Spinocerebellar Ataxia Type 3. Mov Disord 2020; 35: 1679–84. 10.1002/mds.28113.

14 Rezende TJR, Petit E, Park YW, et al. Sensitivity of Advanced Magnetic Resonance Imaging to Progression over Six Months in Early Spinocerebellar Ataxia. Mov Disord 2024. 10.1002/mds.29934.

15 Oliveira CM de, Leotti VB, Polita S, et al. The longitudinal progression of MRI changes in pre-ataxic carriers of SCA3/MJD. J Neurol 2023; 270: 4276–87. 10.1007/s00415-023-11763-6.

16 Faber J, Berger M, Wilke C, et al. Stage-Dependent Biomarker Changes in Spinocerebellar Ataxia Type 3. Ann. Neurol. 2024; 95: 400–06. 10.1002/ana.26824.

17 Schmitz-Hubsch T, Du Montcel ST, Baliko L, et al. Scale for the assessment and rating of ataxia: development of a new clinical scale. Neurology 2006; 66: 1717–20.

18 Jacobi H, Reetz K, Du Montcel ST, et al. Biological and clinical characteristics of individuals at risk for spinocerebellar ataxia types 1, 2, 3, and 6 in the longitudinal RISCA study: analysis of baseline data. Lancet Neurol. 2013; 12: 650–58.

19 Klockgether T, Lüdtke R, Kramer B, et al. The natural history of degenerative ataxia: a retrospective study in 466 patients. Brain 1998; 121: 589–600.

20 Tezenas du MS, Durr A, Rakowicz M, et al. Prediction of the age at onset in spinocerebellar ataxia type 1, 2, 3 and 6. J. Med. Genet. 2014; 51: 479–86.

21 Krismer F, Seppi K, Jönsson L, et al. Sensitivity to Change and Patient-Centricity of the Unified Multiple System Atrophy Rating Scale Items: A Data-Driven Analysis. Mov Disord 2022; 37: 1425–31. 10.1002/mds.28993.

22 Jacobi H, Du Montcel ST, Romanzetti S, et al. Conversion of individuals at risk for spinocerebellar ataxia types 1, 2, 3, and 6 to manifest ataxia (RISCA): a longitudinal cohort study. Lancet Neurol. 2020; 19: 738–47.

23 Grobe-Einsler M, Taheri AA, Faber J, et al. Development of SARA(home), a New Video-Based Tool for the Assessment of Ataxia at Home. Mov Disord. 2021.

24 Chandrasekaran J, Petit E, Park YW, et al. Clinically Meaningful Magnetic Resonance Endpoints Sensitive to Preataxic Spinocerebellar Ataxia Types 1 and 3. Ann. Neurol. 2023; 93: 686–701. 10.1002/ana.26573.

25 Haas E, Incebacak RD, Hentrich T, et al. A Novel SCA3 Knock-in Mouse Model Mimics the Human SCA3 Disease Phenotype Including Neuropathological, Behavioral, and Transcriptional Abnormalities Especially in Oligodendrocytes. Mol. Neurobiol. 2022; 59: 495–522. 10.1007/s12035-021-02610-8.

26 Schuster KH, Zalon AJ, Zhang H, et al. Impaired Oligodendrocyte Maturation Is an Early Feature in SCA3 Disease Pathogenesis. J. Neurosci. 2022; 42: 1604–17. 10.1523/JNEUROSCI.1954-20.2021.

27 Jardim LB, Hauser L, Kieling C, et al. Progression rate of neurological deficits in a 10-year cohort of SCA3 patients. Cerebellum. 2010; 9: 419–28.

28 Jacobi H, Du Montcel ST, Bauer P, et al. Long-term disease progression in spinocerebellar ataxia types 1, 2, 3, and 6: a longitudinal cohort study. Lancet Neurol. 2015; 14: 1101–08.

29 Lin Y-C, Lee Y-C, Hsu T-Y, Liao Y-C, Soong B-W. Comparable progression of spinocerebellar ataxias between Caucasians and Chinese. Parkinsonism. Relat Disord. 2019; 62: 156–62. 10.1016/j.parkreldis.2018.12.023.

30 Peng L, Peng Y, Chen Z, et al. The progression rate of spinocerebellar ataxia type 3 varies with disease stage. J Transl Med 2022; 20: 226. 10.1186/s12967-022-03428-1.

## Appendix References

1. Henschel L, Kugler D, Reuter M. FastSurferVINN: Building resolution-independence into deep learning segmentation methods-A solution for HighRes brain MRI. Neuroimage. 2022;251:118933.

2. Iglesias JE, Van Leemput K, Bhatt P, et al. Bayesian segmentation of brainstem structures in MRI. Neuroimage. 2015;113:184–195.

3. Faber J, Kugler D, Bahrami E, et al. CerebNet: A fast and reliable deep-learning pipeline for detailed cerebellum sub-segmentation. Neuroimage. 2022;264:119703.

4. Andersson JLR, Sotiropoulos SN. An integrated approach to correction for off-resonance effects and subject movement in diffusion MR imaging. Neuroimage. 2016;125:1063–1078.

5. Andersson JL, Skare S, Ashburner J. How to correct susceptibility distortions in spin-echo echo-planar images: application to diffusion tensor imaging. Neuroimage. 2003;20(2):870–888.

6. Tournier JD, Smith R, Raffelt D, et al. MRtrix3: A fast, flexible and open software framework for medical image processing and visualisation. Neuroimage. 2019;202:116137.

7. Fortin JP, Cullen N, Sheline YI, et al. Harmonization of cortical thickness measurements across scanners and sites. Neuroimage. 2018;167:104–120.

8. Fortin JP, Parker D, Tunc B, et al. Harmonization of multi-site diffusion tensor imaging data. Neuroimage. 2017;161:149–170.

9. Faber J, Berger M, Wilke C, et al. Stage-Dependent Biomarker Changes in Spinocerebellar Ataxia Type 3. Ann Neurol. 2024;95(2):400–406.

10. Jacobi H, Reetz K, du Montcel ST, et al. Biological and clinical characteristics of individuals at risk for spinocerebellar ataxia types 1, 2, 3, and 6 in the longitudinal RISCA study: analysis of baseline data. Lancet Neurol. 2013;12(7):650–658.

11. Schmitz-Hubsch T, du Montcel ST, Baliko L, et al. Scale for the assessment and rating of ataxia: development of a new clinical scale. Neurology. 2006;66(11):1717–1720.

